# Dengue risk perception and public preferences for vector control in Italy and France: utility and regret-based choice experiments

**DOI:** 10.64898/2026.04.10.26350604

**Authors:** Filippo Andrei, Michele Tizzoni, Giuseppe Alessandro Veltri

## Abstract

**Background:** Dengue is rapidly emerging in parts of Europe. How households value vector control attributes, and whether inferences depend on decision models or message framing, is unclear.

**Methods:** We conducted a split-ballot online experiment among adults in Italy and France, as well as a hotspot subsample from Marche, Italy. National samples included 1,505 respondents in Italy and 1,501 in France; 183 respondents were recruited in Marche. Participants were randomised to a discrete choice experiment (random utility maximisation) or a regret-based choice experiment (random regret minimisation) and to one of three pre-task messages (control, loss aversion, community values). Each respondent completed 12 choice tasks comparing two dengue control programmes and an opt-out. We estimated mixed logit and mixed random-regret models with random parameters and treatment effects.

**Results:** Across frameworks, nearby cases and high mosquito prevalence were the dominant drivers of programme uptake, whereas cost and operational burden were secondary. In pooled analyses, loss-aversion messaging increased the weight on high mosquito prevalence in both models (from 0.483 to 0.547 in the utility model; from 0.478 to 0.557 in the regret model). Cost effects were small nationally but larger in the hotspot subsample.

**Conclusions:** Risk salience dominates preferences for dengue vector control in these European settings. Random utility and random regret models yield consistent rankings of attributes but differ in behavioural interpretation and some secondary effects; messaging effects were modest and context dependent.

**Key messages:**

- Public preferences for dengue vector control are primarily driven by perceived epidemiological risk, especially proximity to cases and mosquito prevalence, rather than cost or inconvenience.
- Random utility and random regret models produce consistent rankings of key attributes, indicating robust findings across behavioural decision-making frameworks.
- Behavioural messaging (e.g., loss aversion or community framing) has only modest and context-dependent effects on preferences compared to the strong influence of local risk context.

## 1 Introduction

Autochthonous dengue transmission has been reported intermittently in Europe in the last decade, especially in France [1–3], Italy [4–7], and Greece [8], and climate suitability for the establishment of *Aedes* vectors is increasing in parts of the continent [9–11]. Although a safe and effective vaccine exists, it is recommended only for use in endemic regions with high transmission, making vector control the primary strategy for limiting viral spread in non-endemic areas. The adoption of preventive measures to control mosquito proliferation strongly depends on public acceptance of interventions and on public perceptions of dengue risk [12]. To effectively inform the public and promote the uptake of preventive measures, it is essential to understand the factors shaping preferences towards prevention practices in the context of vector-borne diseases, particularly dengue.

In Europe, public awareness of dengue and its associated risks has been investigated by means of traditional surveys [13], typically focusing on small population samples and specific at-risk groups, such as residents of regions with reported local transmission [14, 15]. Previous surveys have usually been developed within the framework of the Health Belief Model, with the aim of measuring knowledge, attitudes, and practices related to the risk of infection by mosquito-borne diseases [16–20]. While standard surveys tend to elicit stated importance or agreement with individual features, discrete choice experiments can quantify how individuals trade off intervention effectiveness, inconvenience, and cost by requiring respondents to choose between realistic alternatives that vary across multiple characteristics [21]. This approach mimics real-world decision-making, allowing estimation of the relative weight placed on each attribute, willingness to pay, and preference heterogeneity. Few studies have adopted choice experiments to investigate the determinants of risk perception of dengue, and most of them have focused on monetary risk attributes in endemic regions [22–24]. Furtheremore, most applications assume random utility maximisation (RUM) [25], but alternative behavioural models, such as random regret minimisation (RRM) [26], conceptualise choices as efforts to avoid the regret of selecting an inferior option, thus offering an alternative framework to interpret individual decisions. Whether these frameworks yield consistent policy-relevant conclusions regarding vector-borne diseases in Europe is unclear.

In this study, we aimed to quantify public preferences for dengue vector control programmes in Italy and France and to test whether brief behavioural messages shift preferences. Using a split-ballot design, we compared inferences from a conventional discrete choice experiment (RUM) and a regret-based choice experiment (RRM). We also oversampled residents of Marche, an Italian region that experienced a large dengue outbreak in 2024 [27], to capture preferences in a higher-risk context where perceived exposure and salience may influence decision-making.

## 2 Methods

### 2.1 Study design and participants

This study employed a split-ballot, online experimental design to investigate public preferences for dengue vector control interventions in Italy and France. The experiment was designed to compare two distinct behavioural frameworks: a discrete choice experiment (DCE) based on the principles of Random Utility Maximisation (RUM), and a regret-based choice (RBC) experiment rooted in the Random Regret Minimisation (RRM) framework.

Participants were first randomised to one of the two experimental groups (DCE or RBC). Within each group, they were further randomised into one of three messaging conditions designed to frame the decision context: a control group that received no preceding message; a loss aversion group exposed to a video message emphasising the negative consequences of inaction; and a community values group that viewed a video highlighting pro-social norms and collective responsibility. This 2 *×* 3 between-subjects factorial design resulted in six experimental groups, allowing for the analysis of both the decision-making framework and the messaging frame on participant choices.

The study population comprised adults aged 18 years and older residing in Italy and France. A stratified sampling strategy was utilised to ensure the final sample was representative of the national population with respect to age, gender, and geographical region. An additional sample of 183 respondents from the Italian region of Marche, where a large dengue outbreak occurred in 2024, was included in the study population. The analytical samples are summarised in Table 1.

**Table 1:** Analytical samples by country and region.

| Variable | Category | N | Percent |
| --- | --- | --- | --- |
| Age | 65 years or older | 1,653 | 27.5 |
|  | 45–54 years | 1,072 | 17.8 |
|  | 55–64 years | 1,022 | 17.0 |
|  | 35–44 years | 942 | 15.7 |
|  | 25–34 years | 778 | 12.9 |
|  | 18–24 years | 547 | 9.1 |
| Country | Italy | 3,011 | 50.1 |
|  | France | 3,003 | 49.9 |
| DCE typology | Standard DCE | 3,008 | 50.0 |
|  | Regret-based DCE | 3,006 | 50.0 |
| Education | High school diploma | 2,107 | 35.0 |
|  | Master’s degree | 1,849 | 30.7 |
|  | Bachelor’s degree | 1,284 | 21.4 |
|  | Primary or lower secondary education | 461 | 7.7 |
|  | PhD | 179 | 3.0 |
|  | Other | 134 | 2.2 |
| Gender | Female | 3,128 | 52.0 |
|  | Male | 2,871 | 47.7 |
|  | Other | 10 | 0.2 |
|  | Prefer not to say | 5 | 0.1 |
| Treatment | Condition B (Loss Aversion) | 2,009 | 33.4 |
|  | Condition C (Community Values) | 2,005 | 33.3 |
|  | Condition A (Control) | 2,000 | 33.3 |
| Tasks per respondent |  | 12 |  |
| Choice tasks | Italy | 36,132 |  |
|  | France | 36,036 |  |

### 2.2 Randomisation and masking

Participants were randomly assigned (1:1) to the random utility (DCE) or the random regret (RBC) arm, and, within each arm, to one of three messaging conditions (control, loss aversion, community values; in the proportion 1:1:1). Allocation was implemented automatically within the survey platform; investigators were not involved in assignment. Because messages and task framing were visible to participants, masking of participants was not feasible.

Ethical approval was obtained from the Research Ethics Board of the University of Trento, document number: 2025-053ESA. All participants provided electronic informed consent before beginning the survey.

### 2.3 Procedures

The experimental procedure was administered via a structured online survey. After providing informed consent, participants progressed through a fixed sequence of modules, as depicted in the Supplementary Information file (Fig. S1). Initially, baseline demographic data were collected, including age, gender, and education level. Subsequently, participants completed a validated 8-item Uncertainty Tolerance Scale to measure their dispositional tolerance for ambiguity. Following this, all participants were presented with standardised educational material detailing the transmission, symptoms, and prevention of dengue fever.

After the educational module, participants were exposed to their assigned video message (or no message for the control group). They were then directed to their assigned choice experiment (DCE or RBC), where they completed 12 sequential choice tasks. Each task required them to choose between two hypothetical dengue control programme scenarios and a status quo option of no intervention. The experiment concluded with a set of follow-up questions assessing the certainty and perceived difficulty of their choices, followed by a full debriefing.

### 2.4 Choice experiment attributes

The scenarios in the choice experiment were defined by six attributes, each with varying levels, as detailed in Table 2. The status quo alternative was fixed at the baseline levels for all attributes (e.g., no cost, low mosquito prevalence). The attribute levels for the hypothetical intervention programmes were varied according to a fractional factorial design optimised for D-efficiency [28].

**Table 2:** Attributes and levels for the discrete choice experiment (DCE and RBC).

| Attribute | Description | Levels |
| --- | --- | --- |
| Cost | One-time household contribution for the intervention | € 0 (covered by the local authority); € 100; € 200 |
| Duration | Duration of the household intervention | 1 hour; 2 hours; 4 hours |
| Disinfestation location | Location of the disinfestation | Inside home; Outside home |
| Home location | Proximity of home to stagnant water | Far from stagnant water; Near stagnant water |
| Mosquito prevalence | Mosquito density in the area | Low; High |
| Proximity to cases | Dengue cases reported in the vicinity | No cases within 100 km; Cases within 50 km |

### 2.5 Statistical analysis

Participant choices were analysed using discrete choice models corresponding to each experimental arm, to quantify attribute trade-offs and test whether messaging frames altered preferences. All models were estimated in R using the Apollo package [29], treating each respondent’s 12 choice tasks as panel data.

#### Discrete Choice Experiment (DCE)

For the DCE, choices were modelled under the RUM framework using a mixed logit (random parameters logit) model. For respondent *i*, alternative *j*, and choice task *t*, utility was specified as

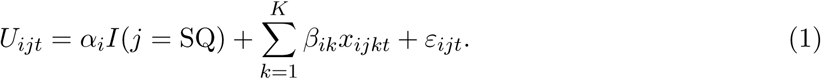

The indicator *I*(*j* = SQ) equals 1 for the opt-out (status quo) alternative and 0 otherwise; thus, a negative ASC*_SQ_* indicates a baseline preference for the intervention alternatives. The error term *ε_ijt_* was assumed independently and identically distributed type-I extreme value, yielding a logit kernel conditional on the individual-specific taste parameters *β_i_*. Preference heterogeneity was captured by specifying the attribute coefficients as random parameters with normally distributed deviations. Messaging treatments were incorporated as mean shifts in each attribute coefficient:

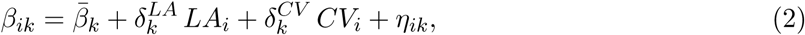

where *LA* and *CV* denote the Loss Aversion and Community Values messaging frames (Control is the reference condition). Continuous attributes (cost and duration) were entered in their natural units, and two-level attributes were represented with binary indicators.

#### Regret-Based Choice (RBC)

For the RBC, choices were analysed using the Random Regret Minimisation (RRM) framework, which assumes that individuals choose the alternative that minimises anticipated regret rather than maximising utility. Regret for choosing alternative *j* in choice task *t* was specified as the sum of attribute-level pairwise comparisons with the other available alternatives:

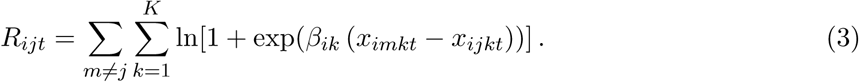

Positive regret weights imply that respondents anticipate more regret when the chosen option performs worse than competing options on a given attribute. The regret weights *β_ik_* were specified as random parameters with the same treatment-specific mean-shift structure as in the DCE, allowing treatment effects to be compared consistently across behavioural frameworks. Choice probabilities follow a logit form in negative regret:

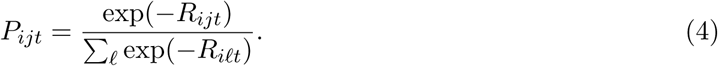

#### Model estimation

The panel likelihood was simulated using 100 Sobol quasi-random draws per respondent, and parameters were estimated via maximum simulated likelihood. All six attribute coefficients were specified as independent normally distributed random parameters. Models were estimated separately for the pooled Italy and France sample, the national Italy sample, the national France sample, and the regional subsample for Marche (Italy), which was identified as a dengue high-risk area.

#### Comparative analysis

Because coefficients from RUM and RRM models are not directly comparable in magnitude (they differ in behavioural interpretation and scale), cross-framework comparisons focused on: (i) the direction and statistical significance of attribute effects; (ii) the rank order and relative importance of attributes, based on the implied change in systematic utility or regret when moving across observed attribute levels; (iii) the propensity to opt out captured by the status quo alternative-specific constant; and (iv) treatment-induced shifts in attribute weights. Results are summarised as treatment-specific mean coefficients with standard errors; 95% confidence intervals are reported for selected estimates in the main text.

## 3 Results

Across both the DCE (RUM) and RBC (RRM) models, attributes related to epidemiological and environmental risk were substantially more influential in shaping participant choices than those related to monetary cost or operational burden. The rank ordering of the key risk attributes was highly consistent across behavioural frameworks, while differences between RUM and RRM were primarily observed for lower-salience operational attributes. The alternative-specific constants for the status quo were negative and statistically significant in all models, indicating a general baseline tendency to choose an active intervention over opting out, conditional on the attributes. The complete set of results is reported in the Supplementary Information (Tables S4 and S5).

### 3.1 Pooled sample (Italy and France)

In the pooled sample, the most important drivers of choice were the proximity of dengue cases and high mosquito prevalence, which consistently showed the largest coefficients in both behavioural frameworks (Figure 1). In the utility model, high mosquito prevalence (vs. low prevalence) increased mean utility in the control group (0.483, 95% CI 0.426–0.540) and was larger under loss aversion (0.547, 95% CI 0.490–0.604). In the regret model, the corresponding regret weights were 0.478 (95% CI 0.402–0.554) in the control group and 0.557 (95% CI 0.481–0.633) under loss aversion. The presence of nearby cases (*≤* 50 km vs *≥* 100 km) was the single most influential attribute in the pooled regret model, 0.777 (95% CI 0.683–0.871) in the control condition. In contrast, intervention cost had a small effect on choices, particularly in the regret model: *−*0.0010 (95% CI *−*0.0020 to *−*0.00002) in the control group.

**Figure 1:**
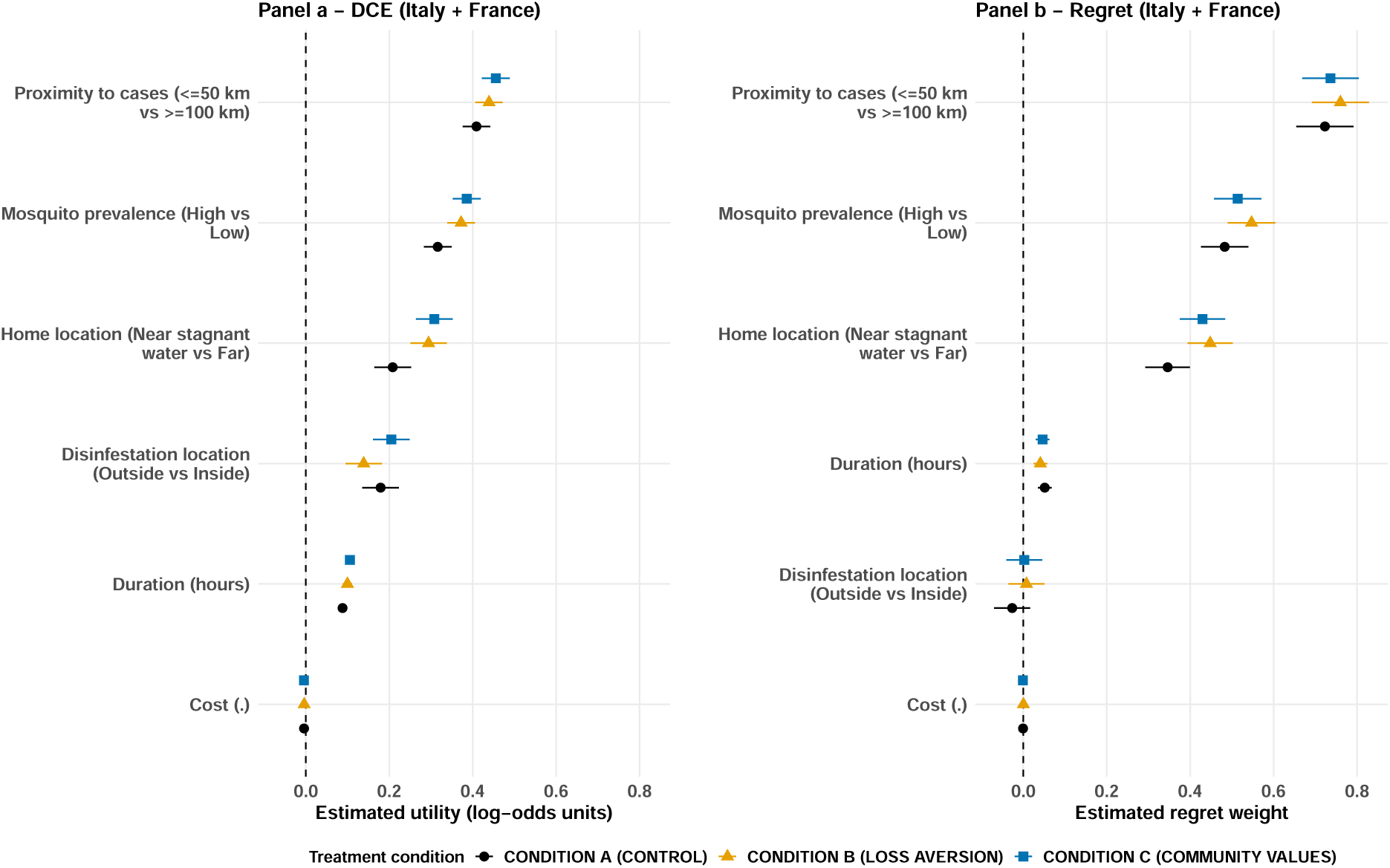
Treatment-specific mean utility and regret weights from the mixed logit and mixed RRM models, pooled Italy and France samples. Points denote mean coefficients by treatment, and horizontal lines show 95% confidence intervals. Symbols denote the treatment condition assigned to the sample: control (circle), loss aversion (triangle), community values (square).

### 3.2 National samples (Italy vs France)

When the analysis was stratified by country, a similar hierarchy of attribute importance was observed, although magnitudes and message effects differed (Figure 2). In Italy, the regret weight for living near stagnant water was 0.435 (95% CI 0.359–0.511) in the control group and increased under loss aversion (0.502, 95% CI 0.426–0.578). In France, the corresponding regret weight was lower in the control group (0.264, 95% CI 0.193–0.335) but increased under both loss aversion (0.369, 95% CI 0.296–0.442) and community values framing (0.365, 95% CI 0.292–0.438). Duration showed a consistent preference for shorter interventions in Italy, whereas in France the estimated duration effect attenuated under community values framing (0.013, 95% CI *−*0.011 to 0.037). Secondary operational attributes showed smaller and more context-specific effects: in Italy, there was a small preference for outdoor disinfestation under loss aversion (0.067, *p <* 0.05), whereas in France there was weak evidence of a preference for indoor disinfestation in the control condition (*−*0.057, *p <* 0.10).

**Figure 2:**
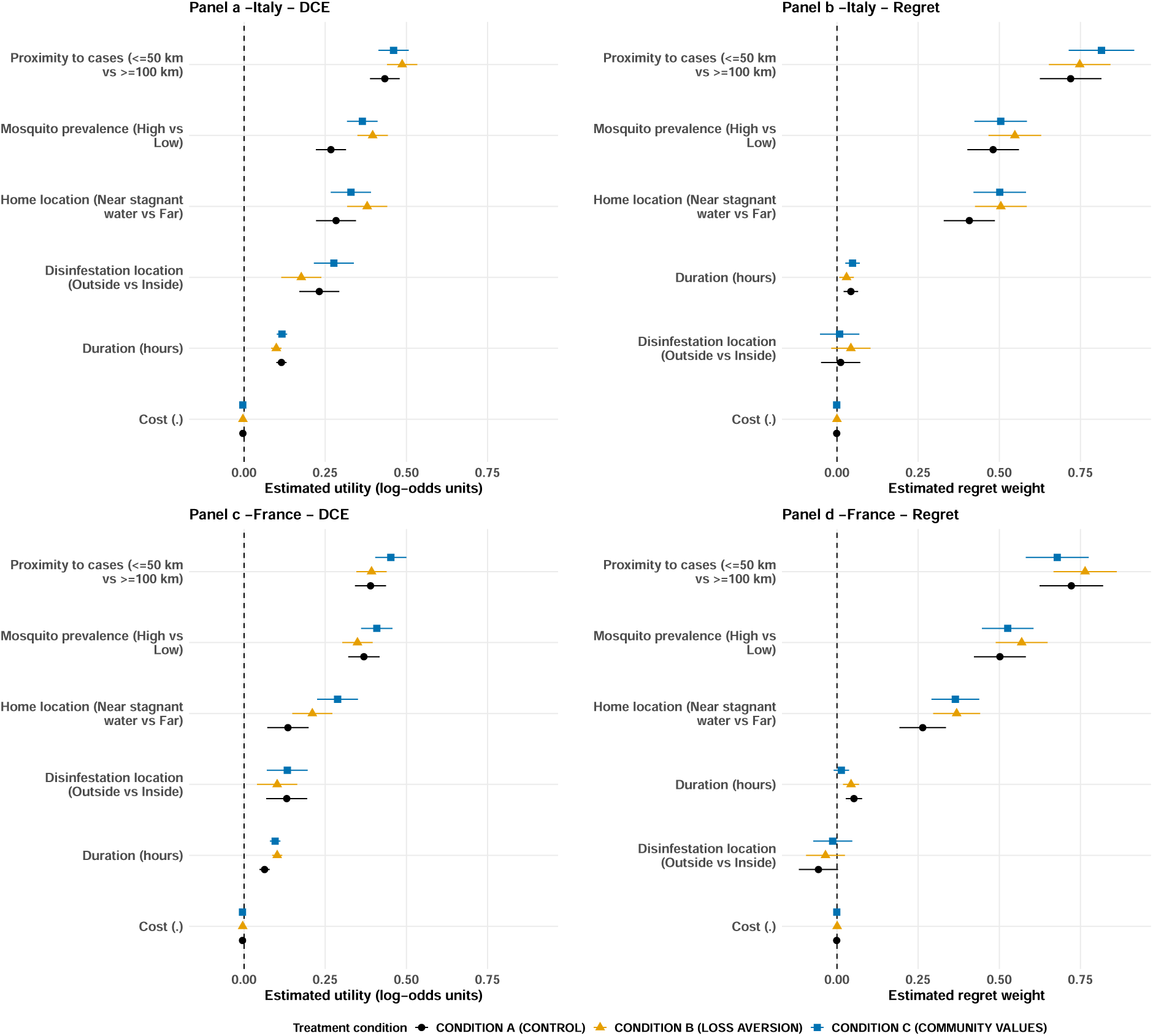
Treatment-specific mean utility and regret weights, stratified by country. Panels a and c show results from the mixed logit (DCE) model for Italy and France, respectively. Panels b and d show results from the mixed RRM model. In all panels, points denote mean coefficients by treatment, and horizontal lines show 95% confidence intervals. Symbols denote the assigned treatment condition: control (circle), loss aversion (triangle), community values (square).

### 3.3 Hotspot subsample (Marche, Italy)

In Marche, an Italian region that experienced a large dengue outbreak in 2024 (see map in the Supplementary Information, Figure S3), sensitivity to risk attributes was particularly pronounced (Figure 3). The regret weight for nearby cases (*≤* 50 km) in the control condition was 1.078 (95% CI 0.782–1.374), substantially higher than in either national sample, indicating heightened risk salience. Unlike the national samples, intervention cost was more salient in Marche (*−*0.0034, *p <* 0.05 in the control group). The baseline preference for an active intervention was also strongest in Marche, as indicated by a large, negative status quo alternative-specific constant (ASC*_SQ_* = *−*1.207, 95% CI *−*1.591 to *−*0.823), implying a low propensity to opt out in this high-risk setting.

**Figure 3:**
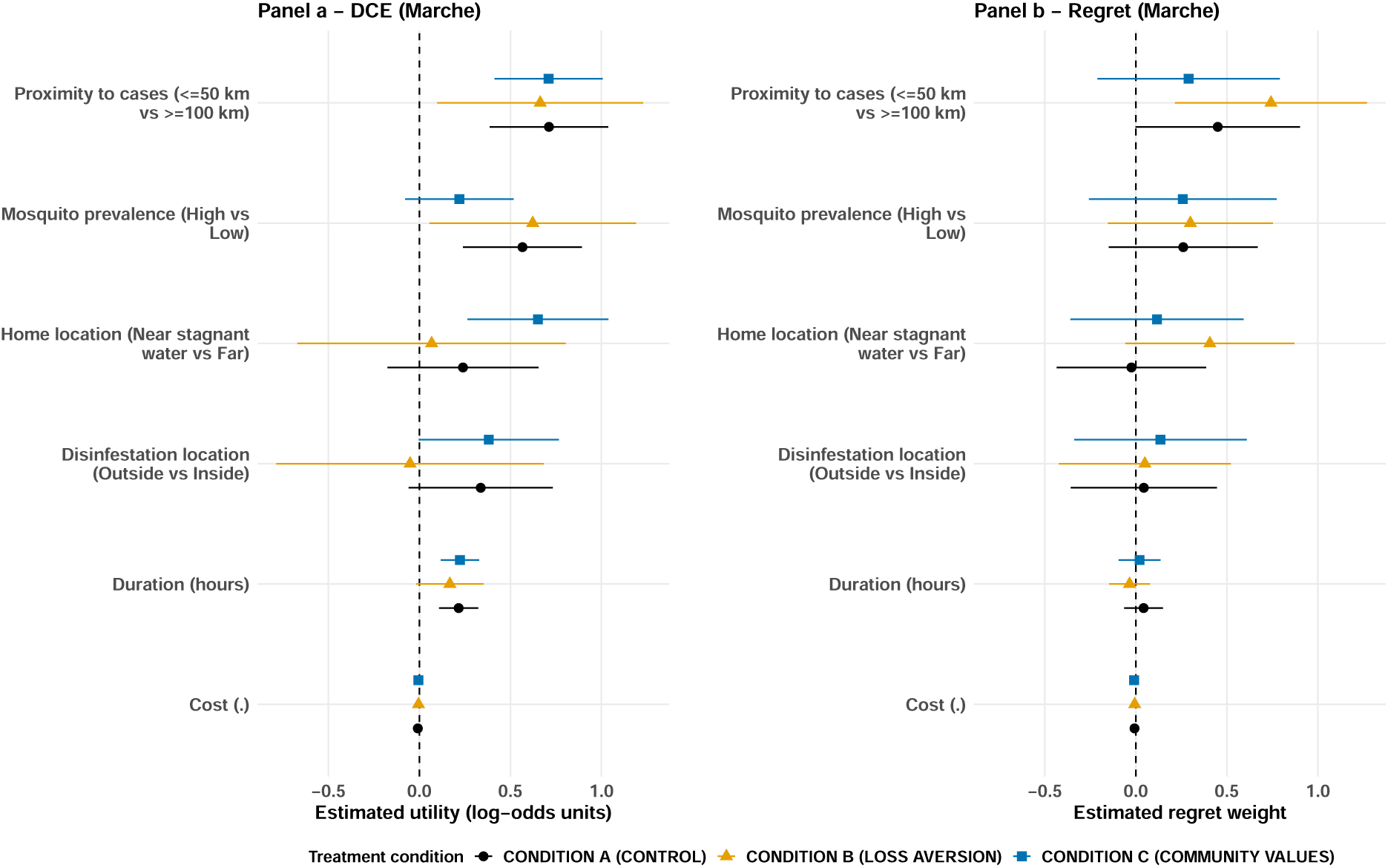
Treatment-specific mean utility and regret weights for the Marche (Italy) subsample. Points denote mean coefficients by treatment, and horizontal lines show 95% confidence intervals. Symbols denote the treatment condition assigned to the sample: control (circle), loss aversion (triangle), community values (square).

### 3.4 Comparison of modelling frameworks and messaging treatments

Across behavioural frameworks, the most prominent finding is the convergence in the hierarchy of preferences: proximity to reported dengue cases and high mosquito prevalence dominate choices in both the RUM and RRM estimations, followed by household proximity to stagnant water. Differences between the two approaches are primarily interpretative—marginal utilities in RUM versus regret weights in RRM—and are most evident for secondary attributes (cost, duration, and disinfestation location), where the magnitude and statistical significance of effects can vary by context. Across both frameworks, treatment effects are modest relative to geographical context: baseline acceptance of intervention (more negative status quo ASCs) is strongest in Marche, intermediate in national Italy, and weakest in France. Messaging frames mainly modulate the weighting of secondary attributes, with the community values frame attenuating duration sensitivity in France and the loss-aversion frame increasing sensitivity to disinfestation location in Italy; in Marche, framing effects are comparatively small relative to the already high salience of risk attributes.

## 4 Discussion

This study provides a comprehensive analysis of public preferences for dengue vector control in two European countries, Italy and France, using a novel experimental design that contrasts two major decision-making frameworks—Random Utility Maximisation (RUM) and Random Regret Minimisation (RRM). Our results consistently demonstrate that attributes related to epidemiological and environmental risk are the most significant drivers of choice, far outweighing concerns about personal cost or the operational burden of interventions. Specifically, the proximity of dengue cases and high mosquito prevalence emerged as the most relevant attributes across all experimental conditions and geographical samples. This indicates that public concern is primarily driven by perceived personal and community risk, a finding with direct implications for public health communication and policy.

A key finding is the heightened risk sensitivity observed in the Marche region of Italy, where a large dengue outbreak occurred in 2024. Residents in this area exhibited substantially higher regret and utility weights for attributes like proximity to cases, and a stronger baseline preference for intervention, suggesting that direct experience with an outbreak significantly amplifies risk perception and demand for protective measures. Furthermore, our analysis revealed that the messaging frames had a modest but discernible impact on preferences. The loss aversion frame, for instance, appeared to increase the salience of certain secondary attributes, such as disinfestation location in Italy, while the community values frame attenuated sensitivity to intervention duration in France. These nuanced effects highlight the context-dependent nature of health messaging.

Our findings align with a growing body of literature that uses stated preference methods to evaluate public awareness and the adoption of vector control measures [23, 24]. Similar to studies conducted in dengue-endemic regions such as Puerto Rico [24] and Cambodia [30], our research confirms that individuals are willing to accept interventions, with preferences strongly shaped by the perceived effectiveness and direct impact on risk exposure. While many studies have focused on willingness-to-pay [31], our work contributes by demonstrating the primary importance of non-monetary risk attributes, a finding that resonates with research showing that factors other than cost are often paramount in health-related decisions [32].

The comparison between RUM and RRM models provides a more nuanced contribution. While some studies in other health contexts have found that RUM models may offer a slightly better fit for aggregate data [33], our results show that both frameworks provide a broadly similar picture of the hierarchy of preferences. The RRM model, however, offers a behaviourally distinct interpretation of choice, suggesting that decisions may be driven by a desire to avoid the negative feeling of regret from making the wrong choice. This is particularly relevant in the context of infectious diseases, where the consequences of inaction can be severe. Our finding that risk attributes carry high regret weights is consistent with the theoretical underpinnings of RRM, which posits that attributes with a larger range of possible outcomes will induce greater potential for regret.

Several psychological mechanisms can explain the strong influence of risk-related attributes. The availability heuristic, where individuals judge the likelihood of an event by the ease with which instances come to mind, likely plays a significant role. For residents in the Marche region, the recent and direct experience of a dengue outbreak makes the risk highly salient and easily imaginable, thus amplifying its importance in decision-making. This is further compounded by dread risk, the heightened fear associated with diseases that are uncontrollable and have severe consequences.

The observed framing effects, although modest, can be understood through the lens of prospect theory. The loss aversion frame [31], which emphasises the negative consequences of not acting, may make the potential losses more salient, leading individuals to be more sensitive to attributes that mitigate those losses. The finding that loss framing can be effective in promoting preventative health behaviours has been documented in other studies, although the underlying mechanism—whether it is true loss aversion or an increase in trust—is still a subject of debate. The community values frame, by contrast, likely works by activating pro-social motivations [34], making individuals consider the collective benefits of their actions. The attenuation of sensitivity to personal burdens like intervention duration in the French sample under this frame suggests a shift from a purely individualistic to a more collectivist decision-making calculus.

Our findings have several important implications for public health policy and communication regarding dengue and other vector-borne diseases in Europe. First, the high importance placed on risk attributes suggests that public health campaigns should prioritise clear and transparent communication about local epidemiological risks. Geographically targeted messaging that provides real-time information on case numbers and mosquito prevalence is likely to be more effective at encouraging uptake of control measures than generic, non-localised advice. For regions with established outbreaks, such as Marche, this is particularly crucial.

Second, the low sensitivity to cost in the national samples suggests that there may be public acceptance for government-funded, rather than individually financed, vector control programmes. While cost became more salient in the hotspot region, risk attributes remained dominant, indicating that even in high-risk scenarios, the primary concern is effectiveness, not price. This provides a strong rationale for public investment in comprehensive vector control strategies.

Third, the nuanced effects of messaging frames suggest that a one-size-fits-all communication strategy is unlikely to be optimal. Health authorities should consider tailoring messages to specific cultural and contextual environments. For instance, in contexts where community cohesion is strong, appeals to collective values may be particularly effective. In others, highlighting the potential personal losses from inaction may be more persuasive. The choice of decision framework (RUM vs. RRM) also has practical implications; if policymakers believe that regret is a key driver of health decisions, then communication strategies could be designed to highlight the potential for post-decisional regret.

Finally, our results could be used to develop computational models of dengue spread that incorporate a behavioural component [32, 35]. For instance, traditional approaches to behavioural epidemic modelling have assumed higher individual awareness to be associated with a higher prevalence of the disease [36]. Our findings provide empirical evidence to support such an assumption when modelling dengue outbreaks in Europe.

There are several limitations to consider when interpreting our findings. First, as a stated preference experiment, the study relies on hypothetical choices. While the results are informative about preferences and trade-offs, actual behaviour may differ. Future research could incorporate real-world payment mechanisms or observe actual uptake of interventions to validate these findings. Second, while our messaging interventions were based on established behavioural theories, the effects were modest. More intensive or prolonged interventions may be needed to produce larger shifts in behaviour. Finally, our analysis focused on two European countries; the findings may not be generalisable to regions with different cultural norms, health systems, or presence of dengue.

In the face of increasing threats from vector-borne diseases in Europe [37], understanding public preferences for control measures is critical for designing effective and acceptable public health strategies. Our study demonstrates that public concern is overwhelmingly driven by the perceived risk of infection, with personal costs and inconveniences playing a secondary role. This suggests that policies should focus on effective risk reduction and that communication should be transparent, localised, and tailored to the specific context. By leveraging insights from behavioural science, public health authorities can build more resilient and responsive systems to fight against the growing challenge of dengue in Europe.

## Ethics approval

Ethical approval for this study was obtained from the Research Ethics Committee of the University of Trento (approval number: 2025-053ESA). The Committee determined, based on an Ethics Self-Assessment, that the study did not involve risks to the psychological or physical well-being of participants, nor did it compromise their rights to privacy, information, or autonomous decision-making. The study involved the processing of personal data and was conducted in accordance with applicable data protection and privacy regulations. All participants provided electronic informed consent prior to participation.

## Author contributions

FA, MT, and GAV contributed to study conceptualisation; FA, MT, and GAV contributed to data collection and methodology; FA and GAV contributed to formal analysis; MT and GAV supervised the study; and FA, MT, and GAV wrote and revised the paper. All authors are responsible for the decision to submit the manuscript.

## Data availability

Data and code are available at the following link: OSF repository https://osf.io/ah75b/overview?view_only=5822f7e3ea0d4cd690558db6d8db90e9.

## Acknowledgments

The authors gratefully acknowledge Alessia Melegaro, Piero Poletti, and Andrea Pugliese for their valuable comments and suggestions.

## Use of artificial intelligence (AI) tools

Artificial intelligence (AI) tools (e.g. ChatGPT) were used to assist with improving the clarity and English language of the manuscript. No AI tools were used in the design of the study, data collection, analysis, or interpretation of results. The authors take full responsibility for the accuracy and integrity of the content.

