## Supplementary Information for "Dengue risk perception and public preferences for vector control in Italy and France: utility and regret-based choice experiments"

### Supplementary appendix

**Figure S1: Experimental study design flowchart**

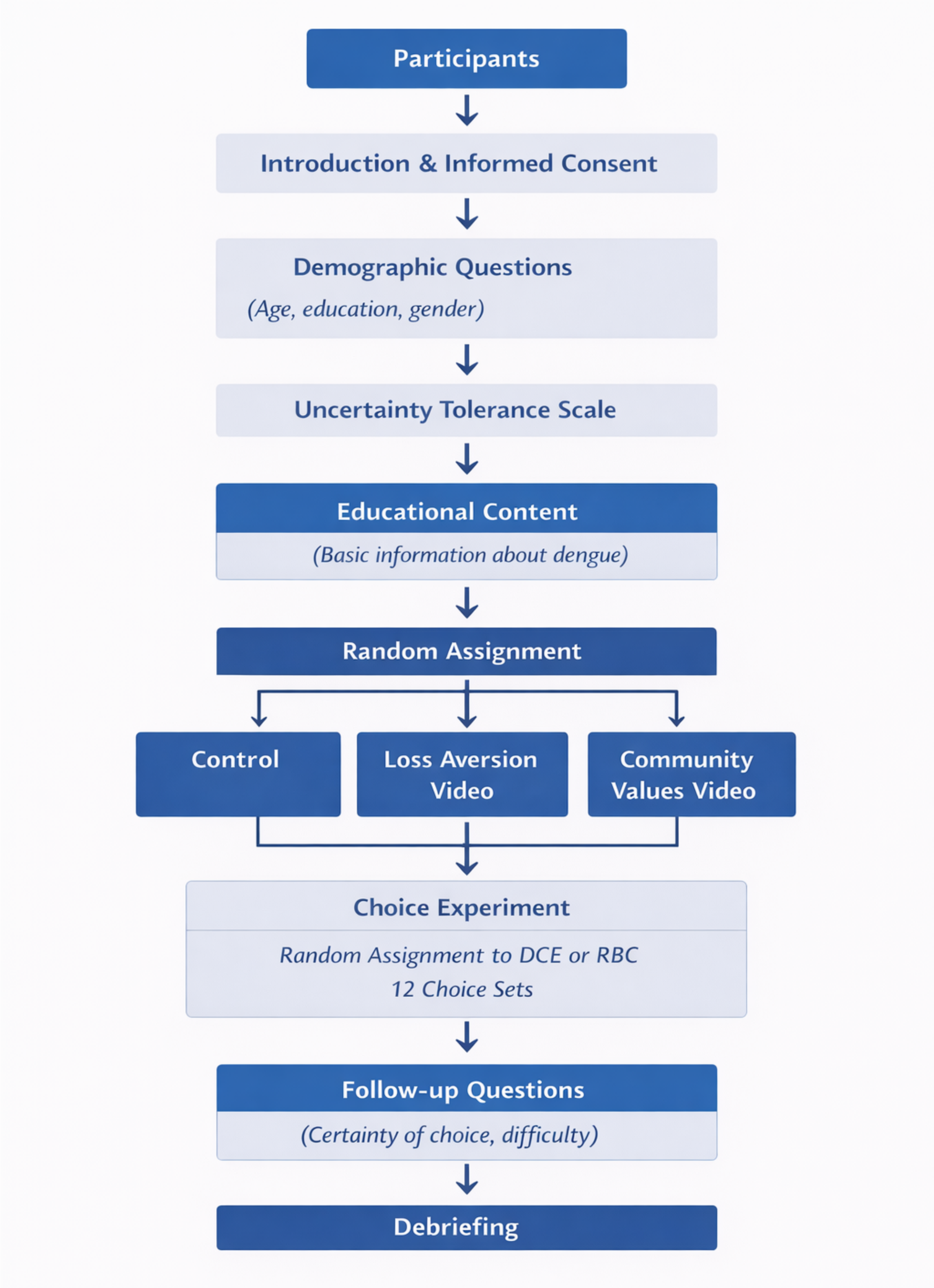

#### SA2: Questionnaire Structure by Phase

**Table 1. Questionnaire Structure by Phase**

| Phase | Component | Description |
|---|---|---|
| 1 | Introduction & Informed Consent | Study description and informed consent |
| 2 | Demographic Information | Age, gender, education |
| 3 | Uncertainty Tolerance Scale | Measurement of tolerance for uncertainty |
| 4 | Educational Content | Essential information about dengue |
| 5 | Random assignment | Control, Loss aversion, Community values video |
| 6 | Practice Choice Set | Example DCE or RBC task |
| 7 | Main Choice Sets | 12 DCE or RBC choice tasks |
| 8 | Follow-up Questions | Certainty, difficulty, attitudes, |
| 9 | Debriefing | Final explanation and contacts |

**PHASE 1: INTRODUCTION AND INFORMED CONSENT**

Welcome

Thank you for agreeing to participate in this study on dengue risk perception and prevention measures.

This survey is conducted at the University of xxx by xxx, xxx, xxx. Your participation is very important for advancing research on the implementation of public health policies. This study, conducted by the University of Trento, aims to better understand how people perceive the risk of dengue and which factors influence prevention-related decisions.

Your participation is completely voluntary and will take approximately 15–20 minutes. Your responses will be processed anonymously and in aggregate form, in full compliance with GDPR regulations.

Informed Consent

☐ I have read and understood the information provided above
☐ I voluntarily agree to participate in this study
☐ I understand that I may withdraw at any time without providing an explanation
☐ I consent to the anonymous processing of my data for research purposes

For any clarification, you may contact:

xxx, xxx, xxx

☐ CONTINUE
☐ EXIT THE STUDY

Consent Declaration

I declare that:

- I have read the information sheet related to the study entitled *"Split Ballot: Dengue Risk Perception and Domestic Access in Europe"*;
- the aims, procedures, and potential risks of the study have been clearly and thoroughly explained to me by xxx xxx xxx

- I had the opportunity to express my views, ask questions, and received satisfactory answers; moreover, I had sufficient time to make an informed, free, and unpressured decision;
- I understand that the data collected have no clinical validity and are intended solely for research purposes;
- I am aware that I may withdraw from the study at any time without providing any explanation;
- The consent I provide to participate in this study is a free decision, not influenced by promises of financial or other benefits, nor by obligations toward the project leaders.

Therefore, being aware of the planned activities and participation procedures:

☐ I GIVE MY CONSENT to participate in the proposed study

☐ I DO NOT GIVE MY CONSENT to participate in the proposed study

**PHASE 2: DEMOGRAPHIC INFORMATION**

**D1. What is your age?**
☐ 18–24
☐ 25–34
☐ 35–44
☐ 45–54
☐ 55–64
☐ 65 or older

**D2. What is your gender?**
☐ Male
☐ Female
☐ Other
☐ Prefer not to say

**D3. Which region do you live in?**
**[**Dropdown menu with Italian/French regions]

D4. What type of area do you live in?
☐ City center
☐ Urban periphery
☐ Suburban area
☐ Rural area
☐ Other

**D5. What type of housing do you live in?**
☐ Apartment building

☐ Detached house with garden

☐ Villa or townhouse

☐ Rural/country house

☐ Other

**D6. What is the highest level of education you have completed?**

☐ Primary or middle school

☐ High school diploma

☐ Bachelor's degree

☐ Master's degree

☐ PhD

☐ Other

**D7. Your home has:**

☐ Air conditioning

☐ Private garden

☐ Balcony/terrace

☐ Courtyard

☐ None of the above

**PHASE 3: TOLERANCE FOR UNCERTAINTY SCALE**

The following statements concern how people deal with uncertain situations.

Please indicate how much you agree with each statement
(1 = Strongly disagree; 5 = Strongly agree):

- TI1. I prefer to have complete information before making a decision
- TI2. I feel comfortable even when I do not know all the details of a situation
- TI3. Unpredictable situations make me anxious
- TI4. I am able to make decisions even when I lack some information
- TI5. I prefer clear and well-defined situations
- TI6. I adapt easily to unexpected changes
- TI7. Uncertainty about the future worries me a lot
- TI8. I am good at handling situations where not everything is clear

Response scale:

☐ 1 = Strongly disagree

☐ 2 = Disagree

☐ 3 = Neither agree nor disagree

☐ 4 = Agree
☐ 5 = Strongly agree

**PHASE 4: EDUCATIONAL CONTENT ON DENGUE**

Information about Dengue

Before proceeding, we would like to share some essential information about dengue.

What is dengue?
Dengue is a viral disease transmitted by mosquitoes of the *Aedes* genus (in particular *Aedes aegypti* and *Aedes albopictus*).

Main symptoms:

- Sudden high fever
- Severe headache
- Muscle and joint pain
- Skin rash
- In severe cases, it may lead to hemorrhagic complications

How it is transmitted:

- Through the bite of infected mosquitoes
- It is NOT transmitted from person to person
- Mosquitoes breed in small collections of stagnant water

Prevention:

- Elimination of mosquito breeding sites
- Use of repellents
- Installation of mosquito nets
- Regular inspection of domestic areas

☐ CONTINUE

**PHASE 5: EXPERIMENTAL MESSAGE**

[NOTE: This section varies depending on the randomly assigned experimental condition]

[CONDITION A – CONTROL]
No message: proceed directly to Phase 5.

[VIDEO – CONDITION B: Negative framing]
[VIDEO – CONDITION C: Social norms and public attention]

Manipulation Check (only for conditions B and C)

Based on the information you heard in the previous video, select the option that best describes the situation:

☐ Online conversations about dengue are mostly accurate and scientific
☐ Online conversations about dengue often contain unverified information
☐ Attention to dengue on social media has increased significantly
☐ I do not remember specific information about social media or online conversations

**PHASE 6: SCENARIO EXAMPLES AND DECISIONS**

The purpose of this part of the study is to understand participants' preferences regarding preventive measures in the event of a dengue outbreak in their area.

Each scenario is based on different attributes such as mosquito prevalence, climate, proximity of cases, type of housing, duration of disinfestation, and cost.

**How the test works**

You will be presented with two scenarios (Scenario A and Scenario B) with different combinations of conditions. For each pair, you will answer two questions:

1. In which scenario would you adopt preventive measures if a dengue outbreak occurred?
2. In which scenario would you consider it more serious not to have taken preventive measures?

Example:

| Attribute | Scenario A | Scenario B |
|---|---|---|
| **Mosquito prevalence** | Low | High |
| **Climate** | Cool and dry | Hot and humid |
| **Proximity of cases** | No cases within 100 km | No cases within 100 km |

| | | |
|---|---|---|
| **Housing** | Rural with stagnant water | Rural, near stagnant water |
| **Disinfestation duration** | 2 hours | 2 hours |
| **Cost** | €100 | €100 |

☐ Scenario A
☐ Scenario B
☐ I would not adopt preventive measures

If a dengue outbreak occurred in your area, in which scenario would you consider it more serious not to have taken preventive measures?

☐ Scenario A
☐ Scenario B
☐ In neither scenario would I consider it serious

**PHASE 7: MAIN CHOICE SETS (DCE OR RBC)**

Participants are randomly assigned to either:

- a Discrete Choice Experiment (DCE) or
- a Risky Binary Choice (RBC) format.

Each participant completes 12 choice sets, constructed using the same structure as the practice example.**Total number of profiles: 24.**

**Task 1**

| Attribute | Option A | Option B |
|---|---|---|
| **Mosquito prevalence** | High | Low |
| **Local climate** | Hot and humid climate | Cool and dry climate |

| | | |
|---|---|---|
| **Proximity of cases** | No cases within 100 km | Cases present within 50 km |
| **Housing conditions** | Modern housing with air conditioning | Rural housing with stagnant water |
| **Disinfestation duration** | 2 hours | 6 hours |
| **Intervention cost** | Covered by the health authority | Covered by the health authority |

**Task 2**

| Attribute | Option A | Option B |
|---|---|---|
| **Mosquito prevalence** | Low | High |
| **Local climate** | Cool and dry climate | Cool and dry climate |
| **Proximity of cases** | Cases present within 50 km | No cases within 100 km |
| **Housing conditions** | Modern housing with air conditioning | Rural housing with stagnant water |

| | | |
|---|---|---|
| **Disinfestation duration** | 4 hours | 6 hours |
| **Intervention cost** | Covered by the health authority | €200 |

**Task 3**

| Attribute | Option A | Option B |
|---|---|---|
| **Mosquito prevalence** | High | Low |
| **Local climate** | Cool and dry climate | Hot and humid climate |
| **Proximity of cases** | Cases present within 50 km | No cases within 100 km |

| | | |
|---|---|---|
| **Housing conditions** | Rural housing with stagnant water | Rural housing with stagnant water |
| **Disinfestation duration** | 2 hours | 6 hours |
| **Intervention cost** | €100 | €200 |

**Task 4**

| Attribute | Option A | Option B |
|---|---|---|
| **Mosquito prevalence** | High | Low |
| **Local climate** | Hot and humid climate | Hot and humid climate |

| | | |
|---|---|---|
| **Proximity of cases** | No cases within 100 km | Cases present within 50 km |
| **Housing conditions** | Rural housing with stagnant water | Modern housing with air conditioning |
| **Disinfestation duration** | 4 hours | 6 hours |
| **Intervention cost** | €100 | €200 |

**Task 5**

| Attribute | Option A | Option B |
|---|---|---|
| **Mosquito prevalence** | Low | High |

| | | |
|---|---|---|
| **Local climate** | Hot and humid climate | Cool and dry climate |
| **Proximity of cases** | No cases within 100 km | Cases present within 50 km |
| **Housing conditions** | Modern housing with air conditioning | Modern housing with air conditioning |
| **Disinfestation duration** | 2 hours | 6 hours |
| **Intervention cost** | Covered by the health authority | €100 |

**Task 6**

| Attribute | Option A | Option B |
|---|---|---|
| **Mosquito prevalence** | Low | High |

| | | |
|---|---|---|
| **Local climate** | Hot and humid climate | Cool and dry climate |
| **Proximity of cases** | Cases present within 50 km | No cases within 100 km |
| **Housing conditions** | Rural housing with stagnant water | Modern housing with air conditioning |
| **Disinfestation duration** | 4 hours | 2 hours |
| **Intervention cost** | €100 | €200 |

**Task 7**

| Attribute | Option A | Option B |
|---|---|---|

| | | |
|---|---|---|
| **Mosquito prevalence** | High | Low |
| **Local climate** | Cool and dry climate | Hot and humid climate |
| **Proximity of cases** | No cases within 100 km | Cases present within 50 km |
| **Housing conditions** | Rural housing with stagnant water | Rural housing with stagnant water |
| **Disinfestation duration** | 6 hours | 2 hours |
| **Intervention cost** | Covered by the health authority | €200 |

**Task 8**

| Attribute | Option A | Option B |
|---|---|---|
| **Mosquito prevalence** | High | Low |
| **Local climate** | Hot and humid climate | Cool and dry climate |
| **Proximity of cases** | Cases present within 50 km | No cases within 100 km |
| **Housing conditions** | Modern housing with air conditioning | Rural housing with stagnant water |
| **Disinfestation duration** | 6 hours | 4 hours |
| **Intervention cost** | Covered by the health authority | €200 |

**Task 9**

| Attribute | Option A | Option B |
|---|---|---|
| **Mosquito prevalence** | Low | High |
| **Local climate** | Hot and humid climate | Cool and dry climate |
| **Proximity of cases** | No cases within 100 km | Cases present within 50 km |
| **Housing conditions** | Modern housing with air conditioning | Modern housing with air conditioning |
| **Disinfestation duration** | 6 hours | 4 hours |
| **Intervention cost** | €100 | €200 |

**Task 10**

| Attribute | Option A | Option B |
| --- | --- | --- |
| **Mosquito prevalence** | High | Low |
| **Local climate** | Hot and humid climate | Cool and dry climate |
| **Proximity of cases** | No cases within 100 km | No cases within 100 km |
| **Housing conditions** | Rural housing with stagnant water | Modern housing with air conditioning |
| **Disinfestation duration** | 4 hours | 2 hours |
| **Intervention cost** | Covered by the health authority | €100 |

**Task 11**

| Attribute | Option A | Option B |
|---|---|---|
| **Mosquito prevalence** | Low | High |
| **Local climate** | Cool and dry climate | Hot and humid climate |
| **Proximity of cases** | Cases present within 50 km | Cases present within 50 km |
| **Housing conditions** | Rural housing with stagnant water | Modern housing with air conditioning |
| **Disinfestation duration** | 2 hours | 4 hours |
| **Intervention cost** | Covered by the health authority | €200 |

**Task 12**

| Attribute | Option A | Option B |
|---|---|---|
| **Mosquito prevalence** | High | Low |
| **Local climate** | Hot and humid climate | Cool and dry climate |
| **Proximity of cases** | Cases present within 50 km | No cases within 100 km |
| **Housing conditions** | Rural housing with stagnant water | Modern housing with air conditioning |
| **Disinfestation duration** | 2 hours | 4 hours |
| **Intervention cost** | €100 | €100 |

**PHASE 8: FOLLOW-UP QUESTIONS**

**F1. How confident are you in the choices you made?**

☐ Not at all confident

☐ Slightly confident

☐ Moderately confident

☐ Quite confident

☐ Very confident

**F2. How did you find the choice task?**

☐ Very difficult

☐ Difficult

☐ Neither easy nor difficult

☐ Easy

☐ Very easy

**F3. Which factors most influenced your decisions? (Select up to 3)**

☐ Mosquito prevalence

☐ Local climate

☐ Proximity to dengue cases

☐ Housing conditions

☐ Disinfestation duration

☐ Cost

☐ Other: ________

**F4. Have you ever had direct or indirect experience with dengue?**

☐ Yes, I or a family member had dengue

☐ Yes, I know someone who had dengue

☐ No, but I had heard about it

☐ No, I had never heard about it before today

**F5. Today, in real life, would you allow a preventive inspection in your home?**

☐ Definitely yes

☐ Probably yes

☐ Not sure

☐ Probably no

☐ Definitely no

**F6. If you answered "Probably no" or "Definitely no," what are your main concerns?**

☐ Privacy/confidentiality

☐ Inconvenience/disturbance

☐ Possible costs

☐ Lack of trust in authorities
☐ I don’t think it’s necessary
☐ Other: ________

**F7. Regardless of your previous answer, which of the following could represent barriers to adopting preventive measures?**
☐ Privacy/confidentiality
☐ Inconvenience/disturbance
☐ Possible costs
☐ Trust in authorities
☐ Perceived necessity
☐ Time required
☐ Required presence during inspection
☐ Other: ________

**PHASE 9: DEBRIEFING**

**Thank you for your participation!**

Your responses have been recorded and will help us better understand how people perceive dengue risk and make prevention decisions.

What we did:

- We studied which factors influence prevention decisions
- There are no right or wrong answers  we were interested in your personal opinions
- Results will help improve public health communication

To learn more about dengue:

- World Health Organization: [link]
- Ministry of Health: [link]
- European Centre for Disease Prevention and Control: [link]

**For questions about this study:**
**xxx**
**xxx**
**xxx**
**xxx**

**Figure S3: Marche Region, Italy, Area of Oversampling**

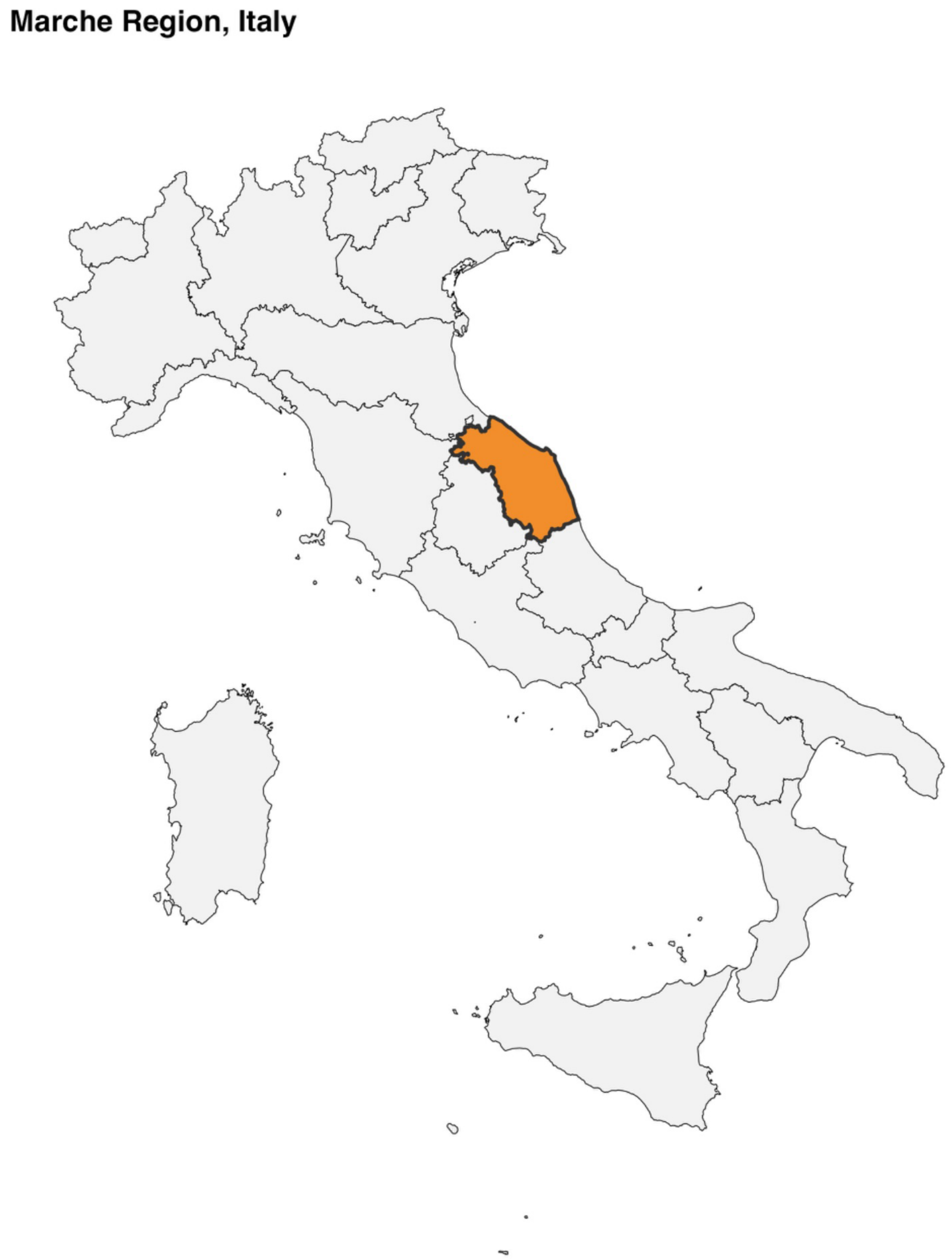

Table S4: **Regret-Based Choice (RBC) Mixed Logit Model Estimates by area**

| **Attribute** | **All** | **Italy** | **French** | **Marche** |
|---|---|---|---|---|
| **Cost (€) Community values** | -0.001** (0.000) | -0.001** (0.001) | -0.001 (0.001) | -0.009*** (0.003) |
| **Cost (€) Control** | -0.001* (0.000) | -0.001* (0.001) | -0.001* (0.001) | -0.007** (0.003) |
| **Cost (€) Loss aversion** | 0.000 (0.000) | -0.000 (0.001) | 0.000 (0.001) | -0.006** (0.003) |
| **Inside (ref.)** | **Reference** | **Reference** | **Reference** | **Reference** |
| **Outside Community values** | 0.002 (0.022) | 0.008 (0.031) | -0.013 (0.031) | 0.135 (0.242) |
| **Outside Control** | -0.027 (0.022) | 0.011 (0.031) | -0.057* (0.031) | 0.044 (0.205) |
| **Outside Loss aversion** | 0.008 (0.022) | 0.043 (0.031) | -0.036 (0.031) | 0.050 (0.241) |
| **Duration (hours) Community values** | 0.046*** (0.008) | 0.048*** (0.011) | 0.013 (0.012) | 0.021 (0.059) |
| **Duration (hours) Control** | 0.051*** (0.008) | 0.043*** (0.011) | 0.052*** (0.013) | 0.043 (0.054) |
| **Duration (hours) Loss aversion** | 0.041*** (0.008) | 0.029*** (0.011) | 0.043*** (0.012) | -0.034 (0.057) |
| **Home far from stagnant water (ref.)** | Reference | Reference | Reference | Reference |
| **Home near stagnant water Community values** | 0.430*** (0.028) | 0.501*** (0.041) | 0.365*** (0.037) | 0.116 (0.243) |
| **Home near stagnant water Control** | 0.346*** (0.027) | 0.408*** (0.040) | 0.264*** (0.036) | -0.024 (0.209) |

| | | | | |
|---|---|---|---|---|
| **Home near stagnant water Loss aversion** | 0.448*** (0.028) | 0.505*** (0.041) | 0.369*** (0.037) | 0.407* (0.237) |
| **Low (ref.)** | Reference | Reference | Reference | Reference |
| **High Community values** | 0.514*** (0.029) | 0.504*** (0.041) | 0.526*** (0.041) | 0.258 (0.263) |
| **High Control** | 0.483*** (0.029) | 0.481*** (0.041) | 0.502*** (0.041) | 0.261 (0.209) |
| **High Loss aversion** | 0.547*** (0.029) | 0.548*** (0.041) | 0.569*** (0.041) | 0.300 (0.231) |
| **No cases within 100 km (ref.)** | Reference | Reference | Reference | Reference |
| **Cases within 50 km Community values** | 0.736*** (0.035) | 0.815*** (0.051) | 0.678*** (0.049) | 0.290 (0.256) |
| **Cases within 50 km Control** | 0.723*** (0.035) | 0.720*** (0.048) | 0.722*** (0.050) | 0.450* (0.230) |
| **Cases within 50 km Loss aversion** | 0.760*** (0.035) | 0.748*** (0.048) | 0.764*** (0.050) | 0.743*** (0.269) |

**Standard errors in parentheses. * p < 0.10, ** p < 0.05, *** p < 0.01.**

**Table S5: Discrete Choice (DCE) Mixed Logit Model Estimates by area**

| Attribute | All | Italy | French | Marche |
|---|---|---|---|---|
| **Cost (€) Community values** | -0.005*** (0.000) | -0.004*** (0.000) | -0.005*** (0.000) | -0.004*** (0.001) |
| **Cost (€) Control** | -0.004*** (0.000) | -0.004*** (0.000) | -0.005*** (0.000) | -0.004*** (0.001) |
| **Cost (€) Loss aversion** | -0.004*** (0.000) | -0.004*** (0.000) | -0.004*** (0.000) | -0.002*** (0.001) |
| **Inside (ref.)** | Reference | Reference | Reference | Reference |
| **Outside Community values** | 0.205*** (0.022) | 0.276*** (0.031) | 0.133*** (0.032) | 0.297*** (0.094) |
| **Outside Control** | 0.179*** (0.022) | 0.232*** (0.031) | 0.131*** (0.032) | 0.246*** (0.091) |
| **Outside Loss aversion** | 0.139*** (0.022) | 0.176*** (0.032) | 0.102*** (0.032) | 0.153 (0.100) |
| **Duration (hours) Community values** | 0.106*** (0.006) | 0.117*** (0.008) | 0.096*** (0.008) | 0.118*** (0.024) |
| **Duration (hours) Control** | 0.088*** (0.006) | 0.115*** (0.008) | 0.063*** (0.008) | 0.152*** (0.023) |
| **Duration (hours) Loss aversion** | 0.100*** (0.006) | 0.099*** (0.008) | 0.102*** (0.008) | 0.129*** (0.026) |
| **Far from stagnant water (ref.)** | Reference | Reference | Reference | Reference |
| **Home near stagnant water Community values** | 0.308*** (0.022) | 0.329*** (0.032) | 0.288*** (0.032) | 0.366*** (0.094) |

| | | | | |
|---|---|---|---|---|
| **Home near stagnant water Control** | 0.208*** (0.022) | 0.283*** (0.031) | 0.135*** (0.032) | 0.321*** (0.091) |
| **Home near stagnant water Loss aversion** | 0.294*** (0.022) | 0.379*** (0.031) | 0.210*** (0.032) | 0.328*** (0.100) |
| **Low (ref.)** | Reference | Reference | Reference | Reference |
| **Mosquito prevalence: High Community values** | 0.545*** (0.024) | 0.515*** (0.034) | 0.578*** (0.035) | 0.460*** (0.102) |
| **Mosquito prevalence: High Control** | 0.447*** (0.024) | 0.378*** (0.033) | 0.522*** (0.035) | 0.395*** (0.098) |
| **Mosquito prevalence: High Loss aversion** | 0.526*** (0.024) | 0.560*** (0.034) | 0.494*** (0.034) | 0.520*** (0.104) |
| **No cases within 100 km (ref.)** | Reference | Reference | Reference | Reference |
| **Cases within 50 km Community values** | 0.644*** (0.024) | 0.651*** (0.034) | 0.639*** (0.035) | 0.780*** (0.101) |
| **Cases within 50 km Control** | 0.578*** (0.024) | 0.613*** (0.033) | 0.551*** (0.034) | 0.843*** (0.097) |
| **Cases within 50 km Loss aversion** | 0.621*** (0.024) | 0.689*** (0.034) | 0.555*** (0.034) | 0.580*** (0.104) |

**Standard errors in parentheses. * p < 0.10, ** p < 0.05, *** p < 0.01.**
